# Perceived Hearing Symptoms Organize the Ear-Disease Comorbidity Network but Are Not a Causal Lever for Brain Health: A Triangulated Analysis

**DOI:** 10.64898/2026.08.13.26360183

**Authors:** Mingyu Chen, Yijia Huang, Ruiqiu Yu, Yan Xie, Feifei Chen, Juezhi Huang, Jinqi Zhao, Zeyu Ma, Zhengrong Ma, Luyun Jiang

**Author notes:** Corresponding author: Luyun Jiang, MD, Chief Physician, Professor, Department of Otolaryngology-Head and Neck Surgery, Affiliated Hospital of Chengdu University of Traditional Chinese Medicine, 39 Shierqiao Road, Jinniu District, Chengdu, Sichuan, China).

## Abstract

**Objectives:** To determine whether objective audiometric hearing loss, subjective hearing difficulty, and tinnitus occupy distinct roles in the population comorbidity network (perceived hub versus audiometric sparse connection), and whether the perceptually active layer exerts an independent causal effect on brain outcomes. Design, Setting, and Participants. Cross-sectional network medicine analysis of 18,939 participants from two nationally representative NHANES cycles (2011-2012, 2015-2016), triangulated with bidirectional Mendelian randomization and external longitudinal validation in CHARLS (China; N = 17,101). Results. Subjective hearing difficulty-not audiometric threshold-acted as the hub linking 16 disease nodes (3 ear, 13 chronic). Bidirectional Mendelian randomization found no robust genetically causal ear-brain association: the null for perceived hearing drew on 22-23 well-identified instruments, whereas objective audiometric loss was indexed by only 3 instruments and was therefore non-informative rather than affirmatively null; across 24 estimable pairs none survived Bonferroni correction (alpha approximately .002). In CHARLS, perceived hearing difficulty predicted incident self-reported memory problems. Conclusions. Perceived hearing symptoms, not audiometric thresholds, organize the ear-disease comorbidity network, yet this perceptually active layer showed no independent causal effect on brain pathology. Subjective ear symptoms are observational flags of multimorbidity-not causal levers-and should prompt comorbidity screening rather than be interpreted as modifiable causal targets.

**Objective:** To test whether objective audiometric hearing loss, subjective difficulty, and tinnitus occupy asymmetric roles in the population comorbidity network (perceived hub vs audiometric sparse connection), and whether the network-active (perceived) layer exerts an independent causal effect on brain outcomes.

**Design, Setting, and Participants:** Cross-sectional network medicine of 18,939 participants in two nationally representative NHANES cycles (2011–2012, 2015–2016), paired with bidirectional MR and external longitudinal validation in CHARLS (China; N = 17,101).

**Exposures:** Sixteen binary disease nodes (3 ear indicators; 13 chronic conditions). MR exposures: objective hearing loss, subjective difficulty, tinnitus.

**Main Outcomes and Measures:** Node degree centrality (network role); bidirectional MR of the ear–brain axis and the reverse direction; incident self-reported memory problems (CHARLS Cox).

**Results:** Bidirectional MR found no well-supported genetically causal ear–brain axis: for perceived hearing the null drew on 22–23 well-identified instruments, whereas objective audiometric loss was represented by only 3 instruments and was therefore non-informative rather than affirmatively null; across 24 estimable pairs, none survived Bonferroni correction (α ≈ .002);

**Conclusions and Relevance:** Perceived hearing symptoms, not audiometric thresholds, organize the ear-disease comorbidity network; yet this network-active layer exerts no independent causal effect on brain pathology. Perceived ear symptoms are flags of multimorbidity, not levers on brain health — arguing for symptom-based, not audiogram-based, screening.

**Key Points:** **Question** Do objective audiometric hearing loss, subjective difficulty, and tinnitus organize the comorbidity network differently, and is there a genetically causal ear–brain axis?

**Findings** In a cross-sectional network of 18,939 NHANES participants, perceived hearing difficulty—not audiometric thresholds—was the hub linking 16 disease nodes; bidirectional Mendelian randomization (24 ear–brain pairs) showed no well-supported genetically causal association.

**Meaning** Subjective hearing symptoms are observational flags, not causal levers, of brain health; they should trigger comorbidity screening rather than be read as modifiable causation.

## 1. Introduction

The 2020 Lancet Commission on Dementia Prevention ranked midlife hearing loss the single largest modifiable dementia risk factor, yet the 2023 ACHIEVE trial—the most definitive hearing-intervention test to date—found no overall cognitive benefit, and even its single positive signal appeared only in a high-cardiovascular-risk subgroup rather than from isolating which hearing construct was treated. This paradox exposes a black box: “hearing loss” is one label for two biologically distinct constructs, audiometric threshold and patient-perceived difficulty^[1,2]^. A patient with a normal audiogram but intrusive tinnitus may carry more anxiety, depression, and vascular comorbidity than one with an elevated threshold and no complaint^[1,2]^—both still called “hearing loss” by threshold alone. Closing this construct gap decides whether hearing care is a causal lever or merely a flag.

Comorbidity network medicine reframes the question^[3,4]^: instead of matching one exposure to one outcome, it maps how diseases co-occur, revealing hubs versus isolates. Centrality describes observed patterns; it does not establish whether a hub drives pathology, reflects reverse causation, or marks shared upstream determinants.

Prior work has treated “hearing loss” as a single, usually audiometric, construct and has not modeled subjective and objective hearing as distinct nodes within one comorbidity architecture, nor triangulated their divergence across bidirectional MR, CHARLS, and LDSC^[5,6]^. We therefore treat the perceptually active layer—subjective difficulty and tinnitus together—as distinct from audiometric threshold, and test whether the two occupy asymmetric network roles.

Having established the perceived layer as the active component, we next test whether it also acts as a causal lever on brain health, triangulating across bidirectional MR, LDSC and two-step MR, and an external CHARLS cohort—independently corroborated by the published ACHIEVE trial^[7]^.

## 2. Methods

### 2.1 Data and population

We analyzed NHANES 2011–2012 and 2015–2016, the two most recent cycles with complete audiometry, tinnitus, and hearing questionnaires. The 2017–2018 cycle was excluded (AUQ191 coding anomaly; audiometry protocol change). After linking AUQ, AUX, DPQ, PFQ, MCQ, DIQ, and BPQ data files, 18,939 participants were included. External validation used CHARLS (China Health and Retirement Longitudinal Study), which enrolled adults aged ≥45 years in 2011, with follow-up through 2018. The analysis plan was preregistered (OSF 9cy4f) before result inspection. The pooled sample spans all ages (median age, 29 y); chronic-condition nodes were questionnaire-defined in adults aged ≥20 y.

### 2.2 Nodes and network

Sixteen binary nodes: 3 ear indicators—objective hearing loss (HL_OBJ; better-ear PTA ≥25 dB), subjective difficulty (HL_SUB; AUQ054), tinnitus (TIN; AUQ191); and 13 cardiometabolic, cerebrovascular, respiratory, and other chronic conditions (eTable 1). Cognitive and dementia constructs were withheld from the node set and analyzed separately as downstream brain outcomes in MR and CHARLS (see §4.4). Edges were pairwise phi coefficients retained at phi 0.10 and P .05 (two-sided); node size ∝ degree centrality; communities by greedy modularity optimization; prevalence by NHANES weights. Encoding the three hearing measures as independent nodes—rather than one “hearing loss” node—quantifies each node’s connectivity separately. The 56.7% audiometry missingness was addressed by repeating the network in complete-case subsample (N = 5,039). Robustness was confirmed via weighted and unweighted sensitivity analyses.

Why the network can separate the two. NHANES measures audiometric threshold and self-reported difficulty with independent items; their missingness differs markedly (objective hearing loss missing in 56.7% of the analytic cohort, versus 0% for self-reported difficulty), so they are not collinear proxies.

### 2.3 Mendelian randomization

Bidirectional 2-sample MR tested the ear-brain axis^[8]^. Exposures: objective hearing loss (ebi-a-GCST90018857), subjective difficulty (ukb-a-257), tinnitus (ukb-b-14254); outcomes: cognition (ebi-a-GCST006572), Alzheimer disease (ieu-a-297; finn-b-G6_ALZHEIMER), dementia (finn-b-F5_DEMENTIA), stroke (ebi-a-GCST005838). Instruments were genome-wide significant SNPs (P 5×10⁻⁸), clumped at r² 0.001/10 Mb, F 10^[9]^; estimation used inverse-variance weighted (IVW) as the primary method, with MR-Egger, weighted median, and weighted mode as secondary methods^[10,11,12]^. The reverse direction tested for reverse causation (OSF Transparent Change A). Full detail in eMethods and eTable 2-3.

### 2.4 Secondary genetic tiers and CHARLS

Three post-registration secondary analyses (OSF Transparent Changes B-D) tested noncausal explanations: LDSC genetic correlation^[13]^, two-step MR through inflammatory mediators, and CAUSE latent-confounder modelling. CHARLS was added post hoc as an external cohort and was not part of pre-registered protocol. In CHARLS, baseline hearing problems (self-reported difficulty, 8.9%) predicted incident self-reported memory problems from 2011 to 2018) using Cox proportional hazards models adjusted for age, sex, and education; Cognitive trajectory was modeled using linear mixed-effects models. Scale-derived impairment cutoffs were excluded; self-reported incident memory problems were the primary outcome.

### 2.5 Ethics

NHANES is public, de-identified, IRB-exempt; CHARLS data were analyzed under a harmonized-data agreement. All genetic analyses used public GWAS summary statistics only (IEU OpenGWAS). Analyses and conclusions are the authors’. Reporting followed STROBE and STROBE-MR^[14]^.

## 3. Results

### 3.1 The “audiometric silence, perceptual activity” paradox (**Figure 1**)

The three ear indicators occupied distinct network roles. Objective hearing loss (degree = 5) connected to subjective difficulty (φ = 0.232) and tinnitus (0.217) within the auditory system and weakly to hypertension (0.128), arthritis (0.125), and diabetes (0.110). Subjective difficulty (degree = 11) bridged hearing into the cardiometabolic-arthritis cluster (eTable 5): arthritis, hypertension, CHD, MI, heart failure, diabetes, stroke, depression, emphysema. Tinnitus (degree = 6) bridged to arthritis, depression, hypertension, bronchitis. Hypertension and arthritis were the highest-degree hubs (degree = 14 each). Perceived symptoms organize multimorbidity; audiometric deficit is sparse and, in adults, absent.

**Figure 1.**
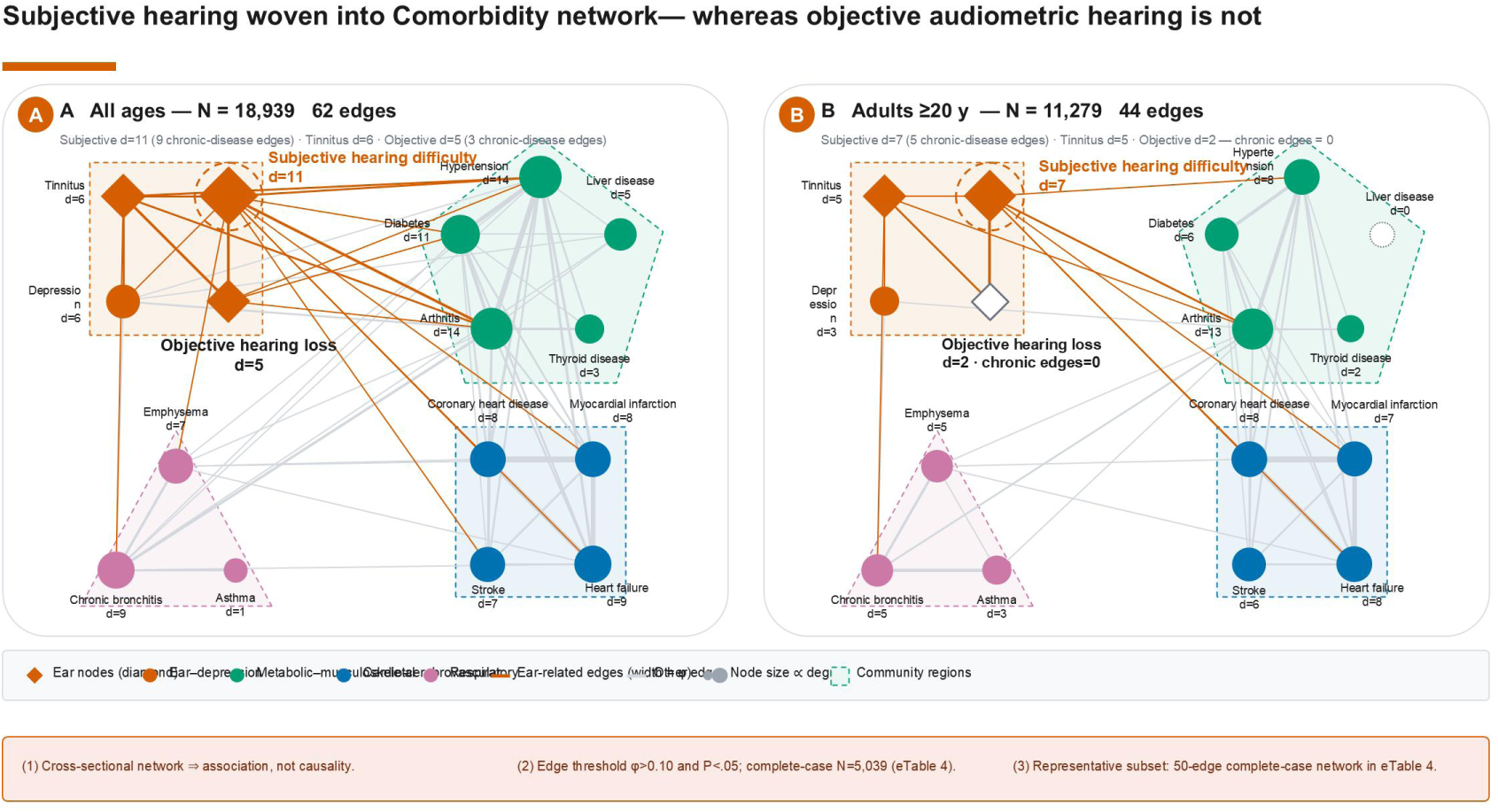
Comorbidity network with independent objective and subjective hearing nodes (two panels). (A) All-ages primary analysis (NHANES 2011–2012 + 2015–2016, N = 18,939, 62 edges); (B) adult sensitivity analysis (≥20 y, N = 11,279, 44 edges). Both panels use a four-column grouped layout by disease system (EAR INDICATORS / CARDIO-METABOLIC / RESPIRATORY / OTHER CHRONIC). Node size ∝ degree; edge line width ∝ phi coefficient (edges retained at phi 0.10, P .05; orange = hearing-related, grey = chronic-disease interconnections). In panel A, subjective hearing difficulty (orange, degree 11) and tinnitus (purple, degree 6) are active hubs spanning cardiometabolic and depressive nodes, whereas objective hearing loss (orange, degree 5) connects to the other two hearing phenotypes and only sparsely to chronic disease. Panel B amplifies the asymmetry: objective hearing loss retains only its hearing-internal edges (degree = 2; no chronic-disease edge), while subjective difficulty remains a hub (degree = 7). Full edge lists in eTable 5 (A) and (B). Cognitive and dementia constructs were intentionally withheld from the node set and analyzed as downstream brain outcomes in the MR and CHARLS streams (Methods; Limitations).

### 3.2 Robustness and communities

The asymmetry persisted in complete-case analysis (N = 5,039), confirming it does not reflect treating missingness as “no disease” (eTable 4): objective hearing loss retained its hearing-internal edges (φ = 0.38 with subjective difficulty) and sparse chronic-disease profile (3 edges); subjective difficulty retained connections to depression, hypertension, and arthritis at reduced degree (5). Weighted and unweighted prevalence were consistent (hypertension 24.0% vs 21.8%). Greedy modularity yielded four communities (Q = 0.16): ear-depression, metabolic-musculoskeletal, cardio-cerebrovascular, and respiratory (eTable 1). Betweenness centrality reproduced the asymmetry (subjective 0.050 vs objective 0.002). Adult-restricted analysis (N = 11,279; 44 edges) confirmed it: objective hearing loss connected only to other hearing phenotypes (degree = 2), subjective difficulty remained a hub (degree = 7) (eTable 6).

We quantified this agreement at the edge level: 48 of 50 complete-case edges (96%) nested within the 62-edge primary network (Jaccard = 0.75; recall = 0.96); the primary analysis recovered 14 additional edges at high-missingness nodes. Zero-imputation thus preserved rather than fabricated structure—consequential given NHANES missingness is structural, not random.

### 3.3 Mendelian randomization: no genetically causal ear-brain axis (**Figure 2**)

We tested whether the perceived-hearing hub exerts an independent causal effect on brain pathology via bidirectional MR. Of 15 planned forward pairs, 9 were estimable: subjective difficulty (5 pairs; 22–23 instruments, F ≈ 30), objective hearing loss (4 pairs; 3 instruments), and tinnitus (0 pairs; 1 instrument). Across all 9, IVW showed no significant effect (all P = .15–.79; eTable 2); none survived Bonferroni (α ≈ .002). MR-Egger intercepts were nonsignificant for all estimable forward pairs (P = .54–.86; objective-HL Egger not estimable with 3 SNPs), and leave-one-out analyses were stable. In reverse, 14 of 15 brain→ear pairs were null; the only nominally significant estimate (cognitive performance → objective HL; β = −0.15; P = .013) was direction-inconsistent across methods—a fragile, pleiotropy-prone signal. Observational centrality does not establish genetic causality.

**Figure 2.**
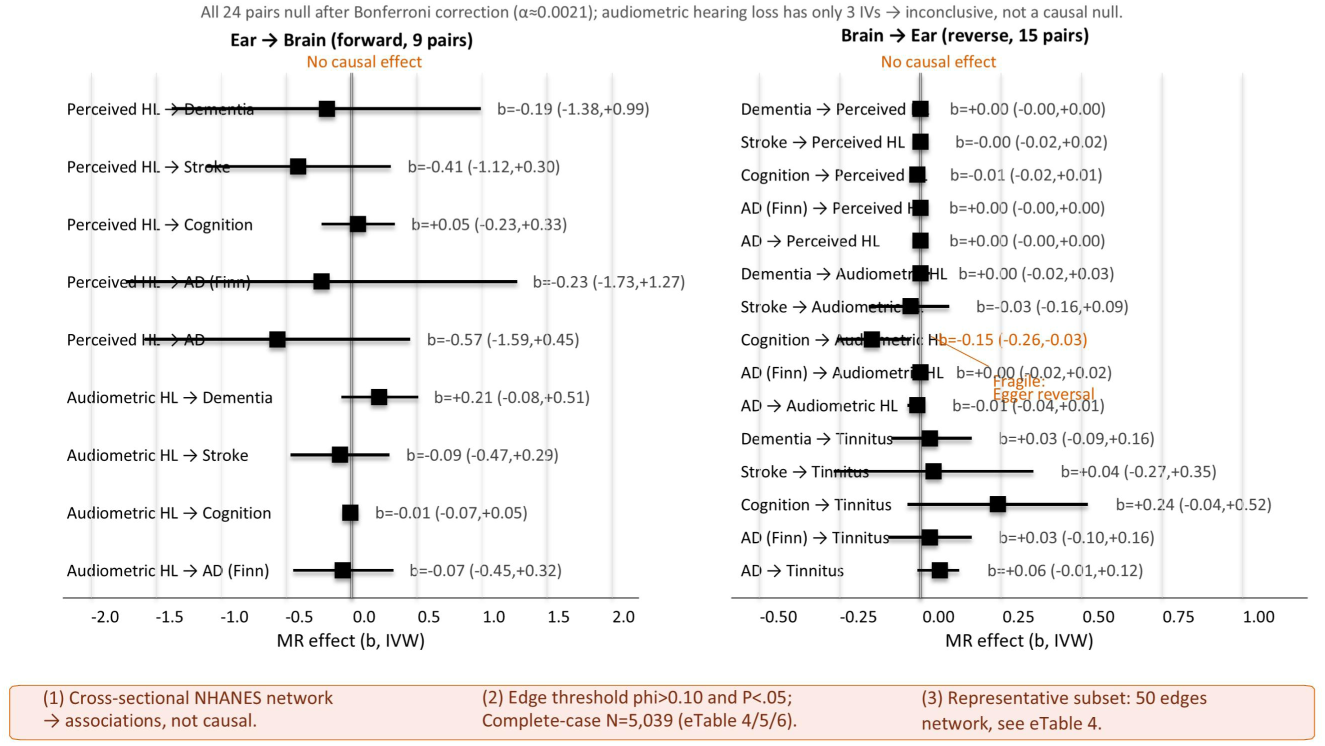
Bidirectional two-sample Mendelian randomization between ear and brain phenotypes (inverse-variance–weighted forest plot; 24 estimable pairs). Forward direction (ear exposure → brain outcome, top) and reverse direction (brain → ear, bottom); all IVW estimates were null after Bonferroni correction (α ≈ .002). Points denote IVW β; bars denote 95% CIs; the vertical dashed line marks the null (β = 0). *Taken together, the MR layer supplies the quantitative basis for ‘observational centrality ≠ genetic causality’: perceived hearing (22–23 instruments) shows no large effect, objective audiometric loss (3 instruments) is noninformative, and the lone reverse signal is direction-inconsistent and pleiotropy-prone. (β units differ by exposure scale; details in eTable 2.)*

### 3.4 Shared genetic architecture and mediation (**Figure 3**)

Tiers 1 and 2 agree with the bidirectional MR: shared genetics and inflammation-mediated pathways are ruled out.

**Figure 3.**
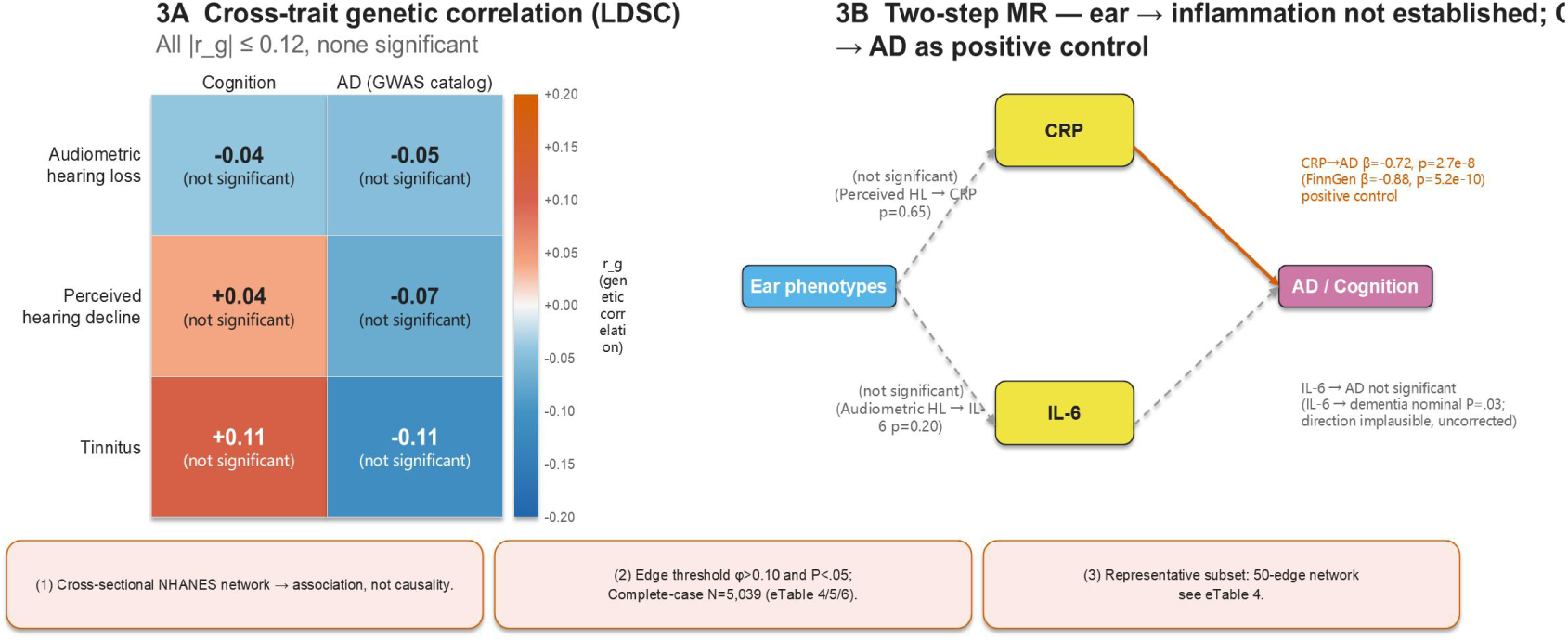
Shared genetic architecture and mediation. Left: LDSC genetic correlation^[13]^ — all 6 estimable ear–brain pairs null (|rg| ≤ 0.12, P = .20–.78). Right: two-step MR — Leg A (inflammation) no mediation (null); Leg B (CRP → Alzheimer disease) significant, reported as a positive control validating the pipeline.

### 3.5 External longitudinal validation in CHARLS (**Figure 4**)

Longitudinal replication (primary external test). Baseline hearing problems (8.9%) predicted incident memory problems (HR = 1.57; 95% CI 1.27–1.94; P = .00004) over 7 years (events = 658), consistent with the network flag.

**Figure 4.**
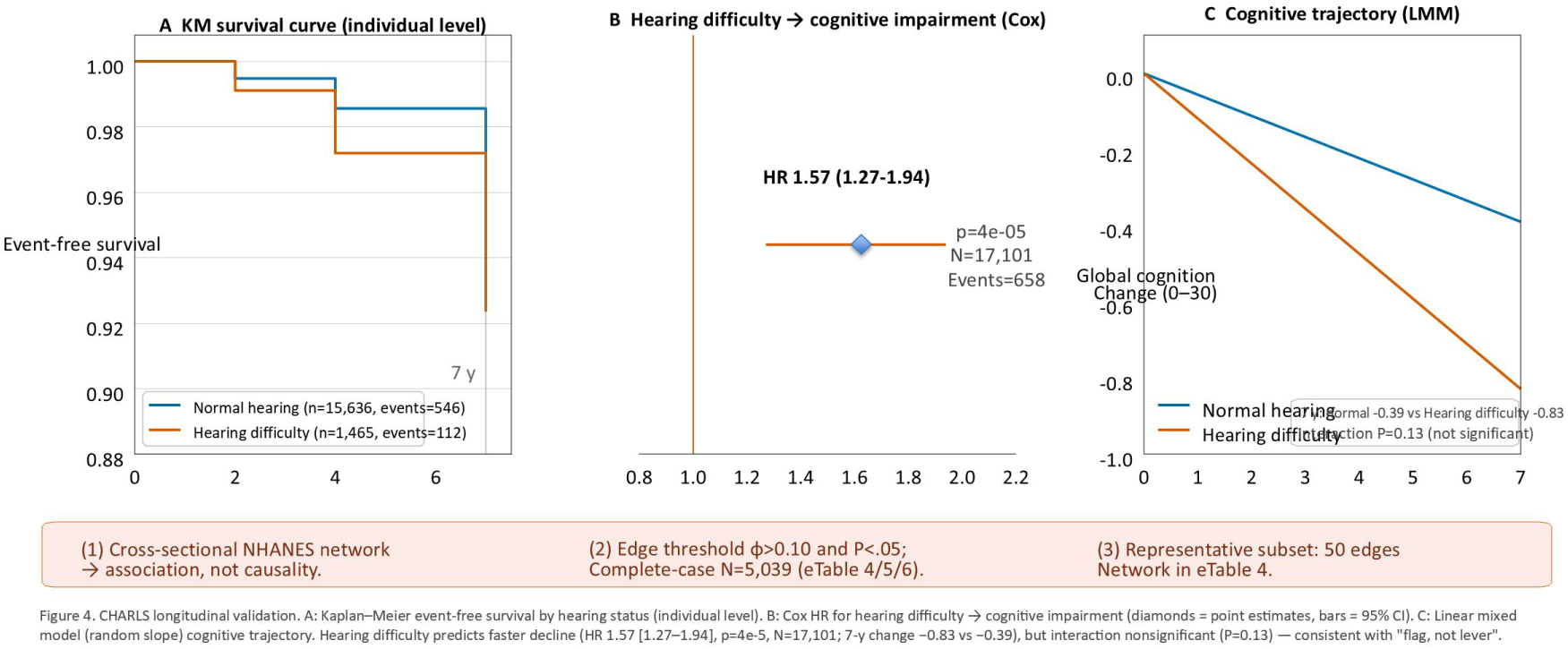
Longitudinal CHARLS: perceived hearing problems predict incident memory problems (Cox model, adjusted for age, sex, education). HR = 1.57 (95% CI 1.27–1.94), P = 4 × 10⁻⁵; N = 17,101; events = 658; 7-year follow-up (2011 baseline → 2018).

**Figure 5.**
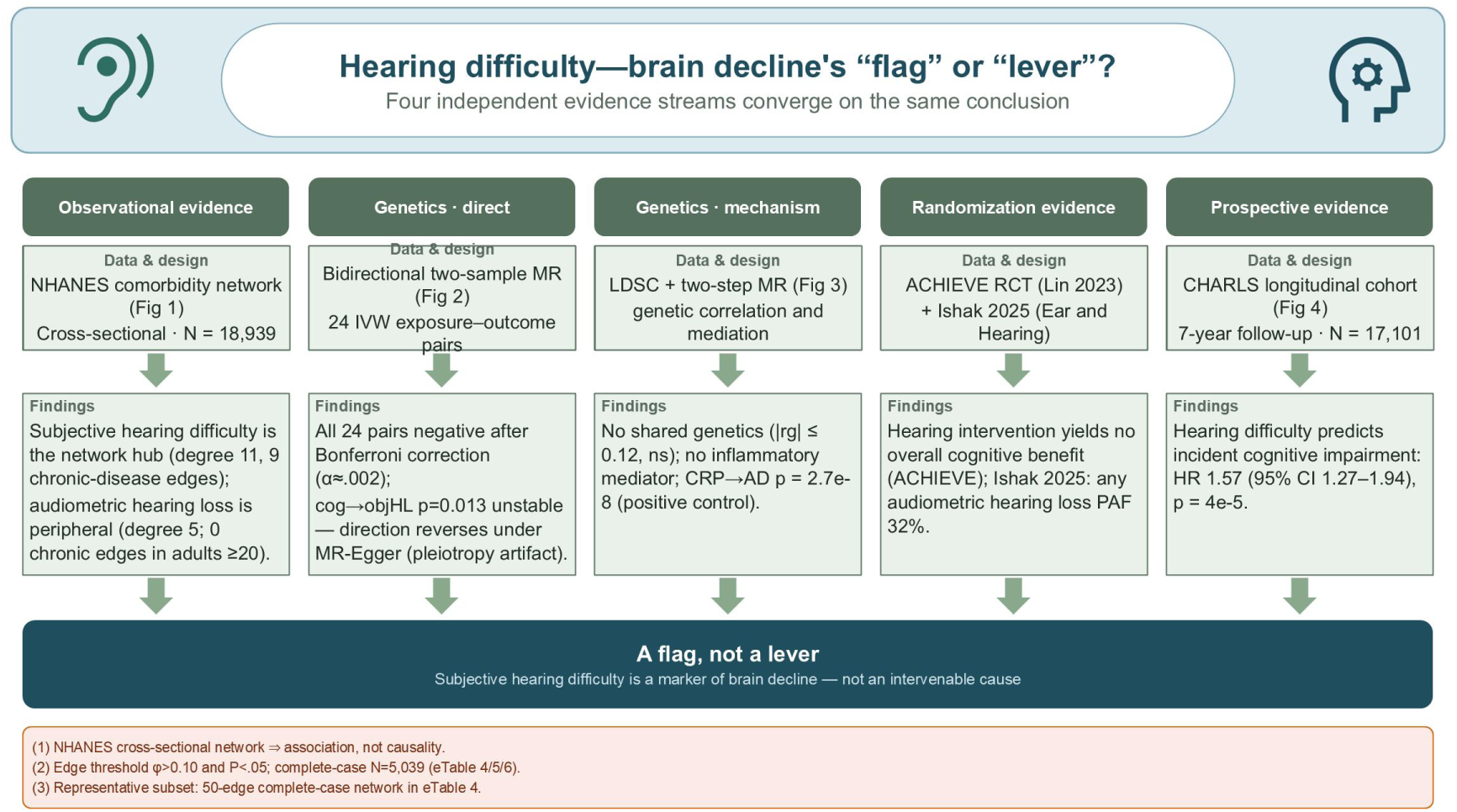
Evidence triangulation across four independent streams. The comorbidity network (perceived hearing as a hub), the longitudinal CHARLS cohort (HR = 1.57), genetic MR (perceived null on 22–23 instruments; objective non-informative on 3 instruments; LDSC |rg| ≤ 0.12), and the randomized ACHIEVE trial (null overall) converge on one inference: perceived hearing symptoms are a flag of multimorbidity, not a causal lever on brain pathology. *One inference stands (Figure 5): perceived hearing is a flag of multimorbidity, not a causal lever—while the objective-audiometry question remains open*.

Exploratory network comparison (not a replication endpoint). In CHARLS 2018 (10 nodes; N = 19,752), subjective hearing difficulty attained degree 0–2 across three definitions—not a failed replication of 11: different node sets (no CHARLS audiometry or tinnitus; cardiac/respiratory nodes collapsed) and the CHARLS schema cap of 7 explain this. Reported as exploratory (Supplement).

Two limits remain regardless of schema. CHARLS collected no audiometry, so the objective-versus-subjective asymmetry could not be tested there; hub centrality is population– and instrument-dependent and need not generalize from an all-ages US sample to a Chinese cohort aged ≥45 y. The longitudinal HR and null MR are unaffected.

## 4. Discussion

### 4.1 Principal findings

This is the first comorbidity network to model objective audiometric and subjective hearing as independent nodes, triangulated with bidirectional MR and CHARLS. The defining result is a two-layer asymmetry—the perceived layer organizes multimorbidity while the audiometric layer is sparse—with no well-supported causal ear–brain axis genetically and perceived hearing predicting incident memory problems longitudinally. Perceived hearing is a risk flag, not a causal lever.

Layer 1 (network). Subjective difficulty (degree = 11) and tinnitus (degree = 6) bridge into the cardiometabolic-arthritis cluster; objective audiometric loss (degree = 5) connects within the auditory system and sparsely to chronic disease (none in adults), an asymmetry not driven by audiometry missingness (§4.4).

Layer 2 (causal). Of 24 estimable MR pairs, 23 were null; the lone nominally significant estimate (cognitive performance → objective HL, P = .013) was direction-inconsistent across methods. The perceived-hearing null drew on 22–23 instruments; objective audiometric loss (3 SNPs) is non-informative (pending). LDSC and two-step MR converged.

Layer 3 (longitudinal, external). In CHARLS (N = 17,101), baseline perceived hearing difficulty predicted incident memory problems over 7 years (HR = 1.57; 95% CI 1.27–1.94; P = .00004), consistent with the network flag; the genetic layer remained null.

### 4.2 The asymmetry is structural, not causal

The divergence is biologically coherent: PTA captures peripheral cochlear damage accumulating silently, whereas subjective difficulty and tinnitus co-occur with depression, social isolation, and functional burden^[2,15,16]^. A composite marker can flag risk better than any single component alone—the ω-6/ω-3 PUFA ratio predicts cognitive decline better than either fatty acid alone^[17]^. The null MR reframes rather than invalidates the network: perceived symptoms remain useful contemporaneous flags, but co-occurrence is not evidence that treating hearing changes brain outcomes.

### 4.3 Clinical implications

The audiogram remains essential for diagnosing cochlear pathology and for aid or implant candidacy, but it is insufficient for flagging multimorbidity risk. The network asymmetry translates into clear clinical and policy implications.

Pathway 1—audiometric deficit without complaint. Auditory rehabilitation is the priority. Treating audiometric loss prevented cognitive decline only in high-risk subgroups (ACHIEVE, 48% reduction[7]) but was null overall; the 32% PAF^[18]^ presumes a causal benefit our null MR does not support, consistent with Labadie’s caution that population attributable fractions (PAF) should not be read as evidence that treating hearing prevents dementia^[19]^.

Pathway 2—difficulty or tinnitus, with or without audiometric loss. The subjective layer is an independent hub of contemporaneous multimorbidity. A brief 2–3-item subjective hearing or tinnitus questionnaire may outperform the audiogram for risk identification and should trigger multimorbidity screening (blood pressure, glucose, inflammation, arthritis, mental health).

Reconciling ACHIEVE with the network. The trial’s 48% slowing in the cardiometabolic-high-risk subgroup is not contrary to “flag, not lever”—it is the expected signal in precisely the multimorbid population our network flags. The null primary outcome and the positive subgroup are therefore consistent: hearing intervention helps selected at-risk individuals, but average audiometric treatment is not a population-level dementia prophylaxis.

Implications for prevention guidelines: The Lancet Commission listed hearing loss as the largest modifiable risk factor for dementia ^[20]^, underscoring its substantial public health weight; our evidence further suggests that, when translating this weight into action, a more prudent framework may not be ‘treat hearing to prevent dementia’ but rather ‘use hearing symptoms as a clue to identify individuals who need cardio-metabolic and neurological preventive care’. This framework both sustains the emphasis on the public health importance of hearing and remains consistent with causal evidence characterizing hearing as a marker rather than a lever.

### 4.4 Limitations

The network is cross-sectional; centrality reflects co-occurrence, not causation. It enumerates physician-diagnosed chronic somatic diseases by design; cognitive and dementia constructs were intentionally withheld from the node set and modeled as downstream brain outcomes in MR and CHARLS, so stroke is the only brain-related node—an explicit scoping decision. Objective-HL missingness (56.7%) is mitigated by the GWAS MR layer; asymmetry persisted in complete cases (N = 5,039). Depression (64.3%) and hypertension (34%) missingness reflect that these interview modules were not administered to all participants (age-eligibility restrictions vary across NHANES components). MR power was asymmetric: subjective HL was well powered (22–23 SNPs, F ≈ 30) and null; objective HL (3 SNPs) and tinnitus (1 SNP) are noninformative and pending; a larger audiometric GWAS is needed. We did not decompose horizontal versus vertical pleiotropy (LDSC and MR-Egger intercepts were null). Generalizability is limited by US sampling and European-ancestry instruments. The asymmetry persisted in adults (≥20 y) and in CHARLS (≥45 y; HR = 1.52; 95% CI 1.22–1.89). Two-step MR interrogated only inflammatory mediators: CRP (264 SNPs) was a valid positive control, but IL-6 carried only 2 instrumental SNPs on the mediator-to-brain leg (underpowered), and vascular mediators (e.g., systolic blood pressure) lacked instruments.

### 4.5 Methodological positioning within the hearing–dementia debate

We do not claim that hearing loss is irrelevant to dementia prevention, nor that the Lancet Commission’s risk ranking is mistaken—observational burden and interventional effect estimate different quantities. The hearing–cognition link is often framed as causal: the 2020 Lancet Commission named hearing loss the largest modifiable dementia risk factor^[20]^, yet ACHIEVE found intervention did not slow decline overall^[7]^. This tension reflects a construct validity gap—audiometric and self-reported hearing are distinct phenotypes^[15]^, and only audiometric loss carries a quantifiable dementia PAF^[18]^—yet MR has treated “hearing loss” as one exposure^[21]^, unable to separate cochlear from perceptual signal. The null MR therefore implies the perceptual layer is a risk flag, not a lever, arguing for symptom-based surveillance rather than hearing treatment as dementia prophylaxis.

The two designs, however, estimate different quantities: ACHIEVE tested 3 years of hearing-aid provision in adults aged 70–84 y, whereas MR estimated lifelong genetic liability—so their results need not agree^[6,22]^. A null genetic estimate does not predict a null trial, nor does a subgroup benefit overturn it.

Our interpretation aligns with a UK Biobank atlas concluding comorbidities reflect shared genetic architecture rather than causality^[5]^ and an earlier analysis that separated objective and self-reported hearing^[23]^. We extend this work by isolating perceived difficulty as an independent hub within a triangulation framework where convergence across unrelated assumptions is the strongest evidence^[6,22]^. A German cohort similarly reports self-reported hearing predicting dementia over 17 years (HR = 1.42)^[24]^.

### 4.6 Conclusion

Patient-perceived auditory symptoms, not audiometric thresholds, organize the ear-disease multimorbidity network yet exert no independent causal effect on brain pathology. Four streams—network (NHANES), longitudinal (CHARLS; HR = 1.57), genetic (null MR), randomized (ACHIEVE)—converge across two populations on shared determinants, not a direct causal chain. Perceived ear symptoms are flags of contemporaneous multimorbidity, not levers on brain health, arguing for symptom-based rather than audiogram-based screening.

## Author contributions

Dr Jiang had full access to all of the data in the study and takes responsibility for the integrity of the data and the accuracy of the data analysis. Concept and design: Chen M, Jiang, Xie. Data acquisition and curation: Chen M, Huang Y, Huang J, Chen F. Statistical and network analysis: Chen M, Yu, Zhao. Mendelian randomization analysis: Yu (methodological design and conduct), Chen M (analysis execution), Xie (methodological review and guidance). Figure and visualization: Chen M, Zhao. Literature search: Huang Y, Chen F, Ma Z. Drafting of the manuscript: Chen M. Critical revision of the manuscript for important intellectual content: Xie,Jiang, Huang Y, Yu, Ma Zh. Supervision, resources, and project administration: Jiang, Xie.

## Conflict of interest disclosures

None reported.

## Data Availability

All data used in this study are publicly available. NHANES data can be accessed at https://www.cdc.gov/nchs/nhanes/. CHARLS data can be accessed at http://charls.pku.edu.cn/ upon registration. Analysis code and network results (canonical JSON files) are provided in the supplementary materials.

https://wwwn.cdc.gov/nchs/nhanes/

http://charls.pku.edu.cn/

https://gwas.mrcieu.ac.uk/

https://www.finngen.fi/en/access_results

https://osf.io/9cy4f/

## Ethical Approval

This study used deidentified public-use NHANES (Centers for Disease Control and Prevention) and CHARLS (Peking University) data. NHANES and CHARLS were approved by their respective institutional review boards, and informed consent was obtained from all participants. Analysis of deidentified secondary data was exempt from additional IRB review at Medical Ethics Committee of the Affiliated Hospital of Chengdu University of Traditional Chinese Medicine (approval No. N/A).

## Acknowledgments

Dr Jiang had full access to all of the data in the study and takes responsibility for the integrity of the data and the accuracy of the data analysis.

Funding/Support: This study was not supported by external funding. Role of the Funder: N/A

Additional Contributions: We thank the NHANES (CDC) and CHARLS participants and investigation teams for the public-use data. STROBE and STROBE-MR reporting checklists are provided in the Supplement. The analysis plan was pre-registered at the Open Science Framework (OSF 9cy4f); post-registration analyses and corrections were logged as Transparent Changes A–F, with a dated erratum covering the metadata of Changes A–D. No compensation was received for this work.

Data Sharing Statement: Deidentified NHANES data are publicly available at https://wwwn.cdc.gov/nchs/nhanes/; CHARLS data are available by application (https://charls.pku.edu.cn/); GWAS summary statistics are publicly available via the IEU OpenGWAS database (https://gwas.mrcieu.ac.uk/). Analysis code, the pre-registered protocol (OSF 9cy4f), and figure-generation scripts are available at https://osf.io/9cy4f/.

## Supplement (eTables)

- eTable 1. Sixteen disease nodes and definitions.
- eTable 2. Forward MR results (IVW, MR-Egger, weighted median, mode).
- eTable 3. Reverse MR results.
- eTable 4. Complete-case robustness (N = 5,039): degree centrality and phi comparison. (subset of the N = 11,279 adult cohort)
- eTable 5. Full network edge list with phi coefficients (N = 62; edges retained at φ 0.10, P .05).
- eTable 6. Adult-restricted (≥20 y) sensitivity: degree centrality comparison (Transparent Change E).
- eTable 7. Network structural descriptors (degree, strength, closeness, betweenness, community assignment; N = 18,939).
- eTable 8. Bidirectional two-sample MR — 24 IVW pairs (ear phenotypes ↔ brain outcomes; Bonferroni α = 0.0021).
- eTable 9. CHARLS longitudinal models — hearing difficulty and cognitive decline (Panel A Cox, Panel B linear mixed model, Panel C Kaplan–Meier).
- eFigure 1. Community-colored network. eFigure 2. Single-cycle cognitive subnet.
- eMethods. Full MR instrument selection, GWAS IDs, LDSC and two-step MR detail, CAUSE specification, STROBE/STROBE-MR checklist.

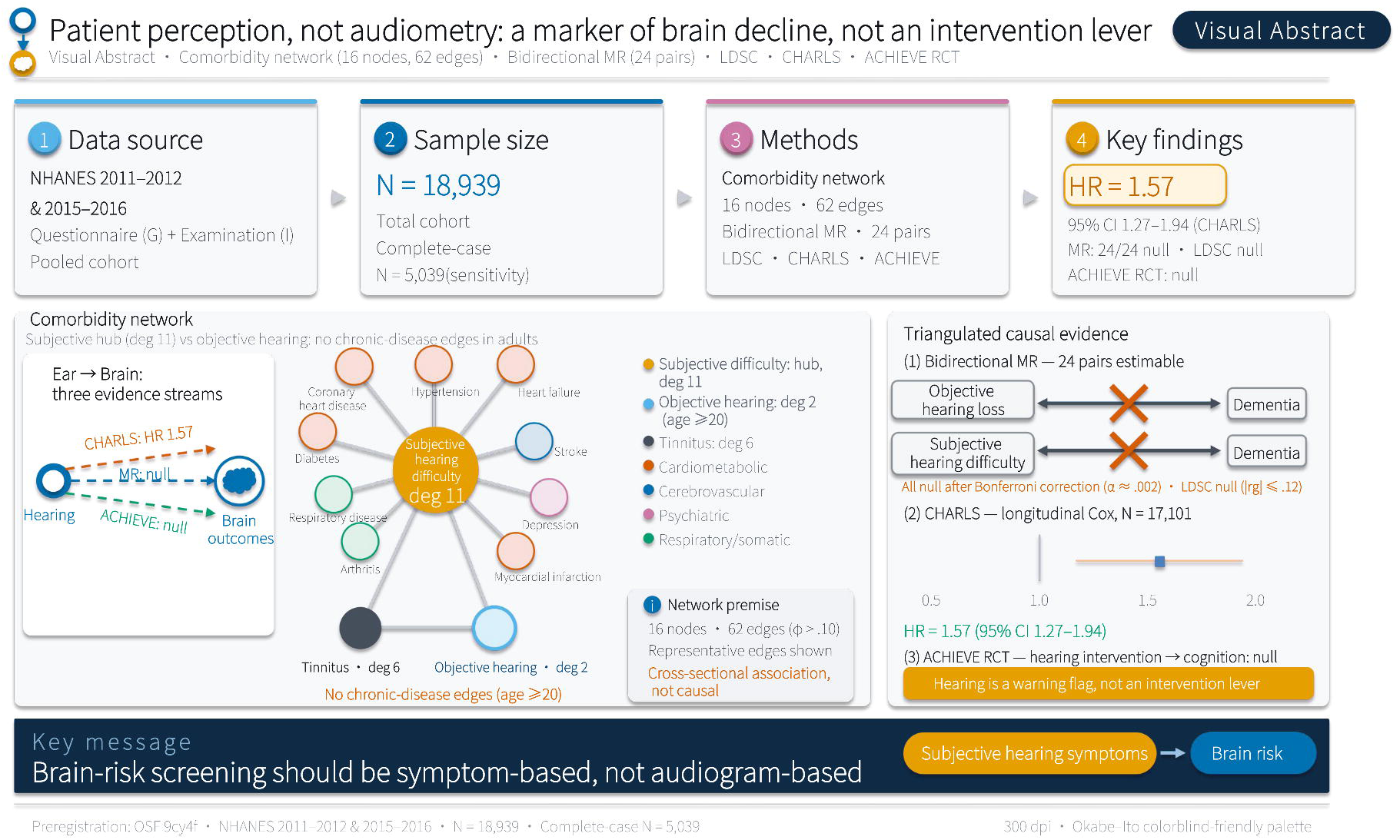

## Notes

### Competing Interest Statement

The authors have declared no competing interest.

### Clinical Protocols

https://osf.io/9cy4f/

### Author Declarations

This study used only publicly available, de-identified datasets that are exempt from institutional review board (IRB) approval: (1) NHANES 2011-2020 from the U.S. CDC/NCHS (https://wwwn.cdc.gov/nchs/nhanes/), a publicly available surveillance dataset with all identifiers removed prior to public release; (2) CHARLS from Peking University (http://charls.pku.edu.cn/), accessed under data use agreement; (3) Genome-wide association summary statistics from OpenGWAS (https://gwas.mrcieu.ac.uk/) and FinnGen (https://www.finngen.fi/en/access_results), which contain only aggregate-level genetic data. All analyses were conducted on de-identified secondary data. The study was determined to be exempt from IRB review by Chengdu University of Traditional Chinese Medicine.

## References

1. Lin FR, Metter EJ, O’Brien RJ, Resnick SM, Zonderman AB, Ferrucci L. Hearing loss and incident dementia. Arch Neurol. 2011;68(2):214-220. doi:10.1001/archneurol.2010.362

2. Mener DJ, Betz J, Genther DJ, Chen D, Lin FR. Hearing loss and depression in older adults. J Am Geriatr Soc. 2013;61(9):1627–1629. doi:10.1111/jgs.12429

3. Barabási AL, Gulbahce N, Loscalzo J. Network medicine: a network-based approach to human disease. Nat Rev Genet. 2011;12(1):56–68. doi:10.1038/nrg2918

4. Van Wijk BCM, Stam CJ, Daffertshofer A. Comparing brain networks of different size and connectivity density using graph theory. PLoS One. 2010;5(10):e13701. doi:10.1371/journal.pone.0013701

5. Bhatt IS, Tucker D, Britton M, et al. A phenome-wide comorbidity atlas of age-related hearing loss, speech-in-noise deficits, and tinnitus: distinguishing causal signals from correlation. J Assoc Res Otolaryngol. 2025;26(6):715–737. doi:10.1007/s10162-025-01008-w

6. Zhang H, Chen K, Gao T, et al. Establishing a robust triangulation framework to explore the relationship between hearing loss and Parkinson’s disease. npj Parkinsons Dis. 2025;11(1):5. doi:10.1038/s41531-024-00861-5

7. Lin FR, Pike JR, Albert MS, et al. Hearing intervention versus health education control to reduce cognitive decline in older adults with hearing loss: the ACHIEVE randomized trial. Lancet. 2023;401(10381):1103–1113. doi:10.1016/S0140-6736(23)01406-X

8. Davies NM, Holmes MV, Davey Smith G. Reading Mendelian randomisation studies: a guide, glossary, and checklist for clinicians. BMJ. 2018;362:k601. doi:10.1136/bmj.k601

9. Burgess S, Thompson SG. Avoiding bias from weak instruments in Mendelian randomization. Int J Epidemiol. 2011;40(3):755–764. doi:10.1093/ije/dyr036

10. Bowden J, Davey Smith G, Haycock PC, Burgess S. Consistent estimation in Mendelian randomization with some invalid instruments using a weighted median estimator. Genet Epidemiol. 2016;40(4):304–314. doi:10.1002/gepi.21965

11. Bowden J, Davey Smith G, Burgess S. Mendelian randomization with invalid instruments: effect estimation and bias detection through Egger regression. Int J Epidemiol. 2015;44(2):512–525. doi:10.1093/ije/dyv080

12. Hartwig FP, Davey Smith G, Bowden J. Robust inference in summary data Mendelian randomization via the zero modal pleiotropy assumption. Int J Epidemiol. 2017;46(6):1985–1998. doi:10.1093/ije/dyx102

13. Bulik-Sullivan BK, Loh PR, Finucane HK, et al. LD Score regression distinguishes confounding from polygenicity in genome-wide association studies. Nat Genet. 2015;47(3):291–295. doi:10.1038/ng.3211

14. Skrivankova VW, Richmond RC, Woolf BAR, et al. Strengthening the reporting of observational studies in epidemiology using Mendelian randomization: the STROBE-MR statement. JAMA. 2021;326(16):1614–1621. doi:10.1001/jama.2021.18236

15. Choi JS, Betz J, Deal J, et al. A comparison of self-report and audiometric measures of hearing and their associations with functional outcomes in older adults. J Aging Health. 2016;28(5):890–910. doi:10.1177/0898264315614006

16. Mick P, Kawachi I, Lin FR. The association between hearing loss and social isolation in older adults. Otolaryngol Head Neck Surg. 2014;150(3):378–384. doi:10.1177/0194599813518021

17. Andrade V, Kleineidam L, Wagner-Thelen H, et al. Prediction of cognitive outcome and progression to dementia using ω6-PUFA/ω3-PUFA ratio. Alzheimers Dement. 2026;22(6):e71590. doi:10.1002/alz.71590

18. Ishak E, Burg EA, Pike JR, et al. Population Attributable Fraction of Incident Dementia Associated With Hearing Loss. JAMA Otolaryngol Head Neck Surg. 2025;151(6):568–576. doi:10.1001/jamaoto.2025.0192

19. Labadie RF. Caution in Interpretation of Population Attributable Fractions. JAMA Otolaryngol Head Neck Surg. 2025;151(10):1010. doi:10.1001/jamaoto.2025.2480

20. Livingston G, Huntley J, Sommerlad A, et al. Dementia prevention, intervention, and care: 2020 report of the Lancet Commission. Lancet. 2020;396(10248):413–446. doi:10.1016/S0140-6736(20)30367-6

21. Jiang D, Hou J, Nan H, et al. Relationship between hearing impairment and dementia and cognitive function: a Mendelian randomization study. Alzheimers Res Ther. 2024;16(1):215. doi:10.1186/s13195-024-01586-6

22. Gutierrez S, Glymour MM, Davey Smith G. Evidence triangulation in health research. Eur J Epidemiol. 2025;40(7):743–757. doi:10.1007/s10654-024-01194-6

23. Xu S, Hou C, Han X, et al. Adverse health consequences of undiagnosed hearing loss at middle age: a prospective cohort study with the UK Biobank. Maturitas. 2023;174:30–38. doi:10.1016/j.maturitas.2023.05.002

24. Stevenson-Hoare J, Stocker H, Trares K, Holleczek B, Brenner H. Subjective hearing and memory problems are associated with dementia and cognition in later life. Alzheimers Dement (Amst). 2024;16(3):e12624. doi:10.1002/dad2.12624

