## Supplementary Material for "Perceived Hearing Symptoms Organize the Ear-Disease Comorbidity Network but Are Not a Causal Lever for Brain Health: A Triangulated Analysis"

### eTable 1. Sixteen Disease Nodes of the Ear-Indexed Comorbidity Network: NHANES Variables, Definitions, Weighted Prevalence, and Centrality

***Abbreviations: PTA, pure-tone average; PHQ-9, Patient Health Questionnaire-9. Two prevalence conventions are reported side by side because 13 of the 16 nodes carry item-level missingness: (i) weighted prevalence among respondents with non-missing data for that item, and (ii) weighted prevalence across the full analytic cohort with missing recoded to 0, which is the convention used to build the network. NHANES sampling weights were applied throughout (WTMEC2YR for audiometric HL_OBJ in convention (i); WTINT2YR for all interview items and for convention (ii); 2-cycle pooling with weights halved). Degree = number of edges at phi > 0.10 and P < .05. Strength = sum of |phi| over a node's edges (edge phi values are listed in eTable 5). Closeness = normalised inverse mean shortest-path distance. Betweenness = Freeman-normalised node betweenness on the unweighted edge set. N = 18,939 (all-ages pooled).***

| Node | NHANES variable | Definition / criterion | Weighted prevalence, non-missing, % | Weighted prevalence, missing = 0, % | Missing, % | Degree | Strength (Σ\|phi\|) | Closeness | Betweenness (Freeman) |
|---|---|---|---|---|---|---|---|---|---|
| **HL_OBJ** | AUXU500/1K/2K/4K | Better-ear 4-frequency pure-tone average (PTA) ≥ 25 dB | 8.0 | 4.7 | 56.7 | 5 | 0.812 | 0.577 | 0.002 |
| **HL_SUB** | AUQ054 | "A little / a lot of trouble hearing" (response 4 or 5) | 6.1 | 6.1 | 0.0 | 11 | 1.653 | 0.750 | 0.050 |
| **TIN** | AUQ191 | Tinnitus present (yes) | 10.6 | 10.6 | 0.0 | 6 | 1.056 | 0.625 | 0.010 |
| **ART** | MCQ160A | Doctor told has arthritis | 24.8 | 18.4 | 40.6 | 14 | 2.757 | 0.938 | 0.136 |
| **CHF** | MCQ160B | Doctor told has congestive heart failure | 2.6 | 1.9 | 40.6 | 9 | 1.855 | 0.714 | 0.018 |
| **DM** | DIQ010 | Doctor told has diabetes | 7.6 | 7.6 | 0.1 | 11 | 1.832 | 0.789 | 0.075 |
| **CHD** | MCQ160C | Doctor told has coronary heart disease | 3.2 | 2.4 | 40.7 | 8 | 1.839 | 0.652 | 0.003 |
| **MI** | MCQ160E | Doctor told had myocardial infarction | 3.3 | 2.4 | 40.5 | 8 | 1.817 | 0.652 | 0.003 |
| **STR** | MCQ160F | Doctor told had a stroke | 2.8 | 2.1 | 40.5 | 7 | 1.100 | 0.625 | 0.000 |
| **EMP** | MCQ160G | Doctor told has emphysema | 1.9 | 1.4 | 40.5 | 7 | 0.966 | 0.652 | 0.010 |
| **BRON** | MCQ160K | Doctor told has chronic bronchitis | 5.7 | 4.3 | 40.6 | 9 | 1.371 | 0.714 | 0.156 |
| **LIV** | MCQ160L | Doctor told has liver disease | 3.7 | 2.7 | 40.5 | 5 | 0.611 | 0.600 | 0.002 |
| **THY** | MCQ160M | Doctor told has thyroid disease | 11.3 | 8.4 | 40.6 | 3 | 0.541 | 0.536 | 0.000 |
| **ASTHMA** | MCQ010 | Ever told had asthma | 15.7 | 15.7 | 0.1 | 1 | 0.188 | 0.429 | 0.000 |
| **DEP** | DPQ020-100 (PHQ-9) | Patient Health Questionnaire-9 total score ≥ 10 | 10.8 | 4.9 | 64.3 | 6 | 0.806 | 0.625 | 0.007 |
| **HTN** | BPQ020 | Doctor told has high blood pressure | 30.2 | 24.0 | 34.1 | 14 | 2.784 | 0.938 | 0.136 |

*Note: Betweenness is Freeman-normalised — divided by (n−1)(n−2)/2 for the 16-node graph — and is computed on the unweighted edge set. The same convention is used in eTable 7 and in Figure 1, so betweenness values are directly comparable throughout the manuscript.*

Subjective hearing difficulty (HL_SUB) is the dominant ear-layer hub: degree 11 (the highest of the three ear nodes, vs HL_OBJ 5 and TIN 6), the highest strength and closeness of the ear layer, and betweenness 0.050. Objective hearing loss (HL_OBJ) sits at the opposite extreme (degree 5, the lowest strength and closeness of the ear layer, betweenness 0.002), quantifying the audiometric-silence / perceptual-activity asymmetry. Centrality was computed on the SEQN-corrected all-ages network (62 edges).

### eTable 2. Forward Mendelian Randomization: Ear Exposure -> Brain Outcome

***Estimates from 2-sample MR (inverse-variance weighted [IVW] primary, with MR-Egger and weighted median). Bonferroni threshold across 24 estimable pairs α ≈ .002. nSNP = number of genome-wide significant instruments ($P < 5×10^{-8}$, $r^2 < 0.001$ within 10 Mb, F > 10). 'Egger p' = MR-Egger intercept P value (directional pleiotropy test); 'Q p' = Cochran Q heterogeneity P value; 'NE' = not estimable (the objective hearing-loss instrument set contains only 3 SNPs, which is insufficient for the Egger intercept). All estimable Egger P values exceed .05, indicating no detectable directional pleiotropy.***

| Exposure | Outcome | nSNP | Method | beta | SE | P | Egger p | Q p |
|---|---|---|---|---|---|---|---|---|
| **objHL** | cog | 3 | IVW | -0.0083 | 0.0314 | 7.9e-01 | NE | 0.744 |
| **objHL** | AD_finn | 3 | IVW | -0.0686 | 0.197 | 7.3e-01 | NE | 0.949 |
| **objHL** | dementia | 3 | IVW | 0.2133 | 0.1497 | 1.5e-01 | NE | 0.776 |
| **objHL** | stroke | 3 | IVW | -0.0868 | 0.1936 | 6.5e-01 | NE | 0.032 |
| **subjHL** | cog | 22 | Egger | 0.1532 | 0.4826 | 7.5e-01 | 0.83 | 0 |
| **subjHL** | cog | 22 | IVW | 0.053 | 0.142 | 7.1e-01 | 0.83 | 0 |
| **subjHL** | cog | 22 | W-med | -0.1267 | 0.1343 | 3.5e-01 | 0.83 | 0 |
| **subjHL** | AD | 22 | Egger | -1.5141 | 2.403 | 5.4e-01 | 0.692 | 0.378 |
| **subjHL** | AD | 22 | IVW | -0.5709 | 0.5223 | 2.7e-01 | 0.692 | 0.378 |
| **subjHL** | AD | 22 | W-med | -0.186 | 0.7124 | 7.9e-01 | 0.692 | 0.378 |
| **subjHL** | AD_finn | 22 | Egger | 1.3952 | 2.7265 | 6.1e-01 | 0.542 | 0.565 |
| **subjHL** | AD_finn | 22 | IVW | -0.2292 | 0.7642 | 7.6e-01 | 0.542 | 0.565 |
| **subjHL** | AD_finn | 22 | W-med | -0.9158 | 1.079 | 4e-01 | 0.542 | 0.565 |
| **subjHL** | dementia | 22 | Egger | 0.1758 | 2.2145 | 9.4e-01 | 0.863 | 0.35 |
| **subjHL** | dementia | 22 | IVW | -0.1948 | 0.6062 | 7.5e-01 | 0.863 | 0.35 |
| **subjHL** | dementia | 22 | W-med | -0.6997 | 0.833 | 4e-01 | 0.863 | 0.35 |
| **subjHL** | stroke | 23 | Egger | -0.9736 | 1.2149 | 4.3e-01 | 0.63 | 0 |
| **subjHL** | stroke | 23 | IVW | -0.407 | 0.3627 | 2.6e-01 | 0.63 | 0 |
| **subjHL** | stroke | 23 | W-med | -0.3521 | 0.3478 | 3.1e-01 | 0.63 | 0 |

Post-hoc power for the objective-hearing-loss -> dementia pair (IVW beta=0.2133, SE=0.1497, OR=1.24, 95% CI 0.92-1.66, P=.15): minimum detectable effect at 80% power = OR >= 1.52; the observed OR=1.24 carried only ~30% power. Thus the null for objective audiometric loss is non-informative (underpowered by 3-SNP precision), not affirmative causal neutrality. Cochran's Q was significant only where multiple methods agreed (subjective HL -> cognition / stroke), supporting a true null rather than a heterogeneity artefact.

### eTable 3. Reverse Mendelian Randomization: Brain Exposure -> Ear Outcome

***Reverse-direction estimates probing reverse causation. 'Egger p' = MR-Egger intercept p-value; 'Q p' = Cochran's Q heterogeneity p-value. The only nominally significant IVW estimate (cognitive performance -> objective HL; P=.01) was direction-inconsistent across methods (see main text) and is classified as a pleiotropy artefact.***

| Exposure | Outcome | nSNP | Method | beta | SE | P | Egger p | Q p |
|---|---|---|---|---|---|---|---|---|
| **cog** | objHL | 146 | Egger | 0.2483 | 0.2736 | 3.7e-01 | 0.142 | 0.05 |
| **cog** | objHL | 146 | IVW | -0.1465 | 0.059 | 1.3e-02 | 0.142 | 0.05 |
| **cog** | objHL | 146 | W-med | -0.0719 | 0.0817 | 3.8e-01 | 0.142 | 0.05 |
| **cog** | subjHL | 146 | Egger | 9e-04 | 0.0285 | 9.8e-01 | 0.776 | 0 |
| **cog** | subjHL | 146 | IVW | -0.007 | 0.0065 | 2.8e-01 | 0.776 | 0 |
| **cog** | subjHL | 146 | W-med | -0.0068 | 0.0078 | 3.8e-01 | 0.776 | 0 |
| **cog** | tinnitus | 146 | Egger | -0.2702 | 0.6241 | 6.7e-01 | 0.4 | 0.011 |
| **cog** | tinnitus | 146 | IVW | 0.2424 | 0.1434 | 9.1e-02 | 0.4 | 0.011 |
| **cog** | tinnitus | 146 | W-med | 0.1535 | 0.1954 | 4.3e-01 | 0.4 | 0.011 |
| **AD** | objHL | 15 | Egger | -0.0156 | 0.0171 | 3.8e-01 | 0.752 | 0.204 |
| **AD** | objHL | 15 | IVW | -0.0121 | 0.0127 | 3.4e-01 | 0.752 | 0.204 |
| **AD** | objHL | 15 | W-med | -0.0156 | 0.0122 | 2e-01 | 0.752 | 0.204 |
| **AD** | subjHL | 14 | Egger | 0.0012 | 0.0017 | 5e-01 | 0.995 | 0.25 |
| **AD** | subjHL | 14 | IVW | 0.0012 | 0.0012 | 3.4e-01 | 0.995 | 0.25 |
| **AD** | subjHL | 14 | W-med | 0.0014 | 0.0011 | 2.4e-01 | 0.995 | 0.25 |
| **AD** | tinnitus | 14 | Egger | 0.0685 | 0.0441 | 1.5e-01 | 0.714 | 0.203 |
| **AD** | tinnitus | 14 | IVW | 0.0579 | 0.0328 | 7.7e-02 | 0.714 | 0.203 |

| | | | | | | | | |
|---|---|---|---|---|---|---|---|---|
| **AD** | tinnitus | 14 | W-med | 0.0617 | 0.0322 | 5.5e-02 | 0.714 | 0.203 |
| **AD_finn** | objHL | 3 | Egger | 0.0191 | 0.0179 | 4.8e-01 | 0.447 | 0.401 |
| **AD_finn** | objHL | 3 | IVW | 0.0027 | 0.0112 | 8.1e-01 | 0.447 | 0.401 |
| **AD_finn** | objHL | 3 | W-med | 0.0051 | 0.0115 | 6.6e-01 | 0.447 | 0.401 |
| **AD_finn** | subjHL | 3 | Egger | 1e-04 | 0.0021 | 9.6e-01 | 0.787 | 0.449 |
| **AD_finn** | subjHL | 3 | IVW | 7e-04 | 0.0011 | 5.2e-01 | 0.787 | 0.449 |
| **AD_finn** | subjHL | 3 | W-med | 8e-04 | 0.0011 | 5e-01 | 0.787 | 0.449 |
| **AD_finn** | tinnitus | 3 | Egger | 0.0507 | 0.1441 | 7.8e-01 | 0.87 | 0.005 |
| **AD_finn** | tinnitus | 3 | IVW | 0.0276 | 0.0663 | 6.8e-01 | 0.87 | 0.005 |
| **AD_finn** | tinnitus | 3 | W-med | 0.042 | 0.0285 | 1.4e-01 | 0.87 | 0.005 |
| **dementia** | objHL | 6 | Egger | 0.0255 | 0.0205 | 2.8e-01 | 0.213 | 0.553 |
| **dementia** | objHL | 6 | IVW | 0.0038 | 0.0143 | 7.9e-01 | 0.213 | 0.553 |
| **dementia** | objHL | 6 | W-med | 0.0127 | 0.0152 | 4.1e-01 | 0.213 | 0.553 |
| **dementia** | subjHL | 6 | Egger | 0.0034 | 0.0025 | 2.4e-01 | 0.326 | 0.169 |
| **dementia** | subjHL | 6 | IVW | 0.0014 | 0.0018 | 4.2e-01 | 0.326 | 0.169 |
| **dementia** | subjHL | 6 | W-med | 0.0026 | 0.0015 | 7.6e-02 | 0.326 | 0.169 |
| **dementia** | tinnitus | 6 | Egger | 0.0471 | 0.0993 | 6.6e-01 | 0.869 | 0.012 |
| **dementia** | tinnitus | 6 | IVW | 0.0348 | 0.0627 | 5.8e-01 | 0.869 | 0.012 |
| **dementia** | tinnitus | 6 | W-med | 0.0411 | 0.0393 | 3e-01 | 0.869 | 0.012 |
| **stroke** | objHL | 17 | Egger | -0.1314 | 0.4118 | 7.5e-01 | 0.815 | 0.132 |
| **stroke** | objHL | 17 | IVW | -0.0345 | 0.0622 | 5.8e-01 | 0.815 | 0.132 |
| **stroke** | objHL | 17 | W-med | -0.0346 | 0.0764 | 6.5e-01 | 0.815 | 0.132 |
| **stroke** | subjHL | 17 | Egger | 0.0988 | 0.0756 | 2.1e-01 | 0.203 | 0 |
| **stroke** | subjHL | 17 | IVW | -6e-04 | 0.0116 | 9.6e-01 | 0.203 | 0 |
| **stroke** | subjHL | 17 | W-med | 0.0138 | 0.0086 | 1.1e-01 | 0.203 | 0 |
| **stroke** | tinnitus | 17 | Egger | 0.7595 | 1.0682 | 4.9e-01 | 0.507 | 0.107 |
| **stroke** | tinnitus | 17 | IVW | 0.0415 | 0.1581 | 7.9e-01 | 0.507 | 0.107 |
| **stroke** | tinnitus | 17 | W-med | 0.1651 | 0.1971 | 4e-01 | 0.507 | 0.107 |

### eTable 4. Complete-Case Robustness (SEQN-correct alignment; N = 5,039)

***Degree centrality under the primary missing-as-0 network (N = 18,939, 62 edges) versus the pre-registered complete-case network, in which all 16 nodes are observed (N = 5,039, 50 edges). Both networks were rebuilt from the SEQN-explicit merge (build_network_v3_seqn.py); the complete-case figure replaces the earlier misaligned N = 9,082 and the superseded N = 5,123 / 52-edge count produced before the SEQN fix. Because objective audiometry is administered only to adults aged 20 years or older, this complete-case sample (N = 5,039) is a subset of the N = 11,279 adult cohort (Figure 1, panel B), not an independent all-ages sample.***

| Node | Primary network degree (N=18,939) | Complete-case degree (N=5,039) | Interpretation |
|---|---|---|---|
| **HL_OBJ** | 5 | 5 | Unchanged; identical partners (2 hearing-internal, 3 chronic) |
| **HL_SUB** | 11 | 5 | Attenuated; retains depression, hypertension, arthritis |
| **TIN** | 6 | 6 | Unchanged; identical partners |
| **ART** | 14 | 15 | Stable hub |
| **CHF** | 9 | 7 | Modest attenuation |
| **DM** | 11 | 8 | Modest attenuation |
| **CHD** | 8 | 7 | Modest attenuation |
| **MI** | 8 | 6 | Modest attenuation |
| **STR** | 7 | 4 | Attenuated |
| **EMP** | 7 | 6 | Modest attenuation |
| **BRON** | 9 | 8 | Modest attenuation |
| **LIV** | 5 | 4 | Modest attenuation |
| **THY** | 3 | 1 | Attenuated (low degree throughout) |
| **ASTHMA** | 1 | 3 | Increased |
| **DEP** | 6 | 4 | Modest attenuation |

| HTN | 14 | 11 | Stable hub |
|---|---|---|---|

Edge-level robustness: the primary missing-as-0 network (N = 18,939; 62 edges) and the complete-case network (N = 5,039; 50 edges) share 48 of the 50 complete-case edges (Jaccard = 0.750; recall = 0.960), so the reported structure is not an artefact of zero-imputation. The primary network additionally recovers 14 edges that the complete-case network is too small to detect. Edge lists: network_allages_v3seqn.json (62 edges) and network_completecase_v3seqn.json (50 edges), both produced by build_network_v3_seqn.py and tabulated in eTable 5.

### eTable 5. Full Edge List of the Ear-Indexed Comorbidity Network (phi > 0.10 and P < .05)

***Every retained edge with its phi coefficient and two-sided P value (chi-square test on 1 df). Panel A is the primary all-ages network and Panel B the adult (≥20 y) sensitivity network; these are Panels A and B of Figure 1. Edges are grouped as hearing-internal (both endpoints in the ear layer), hearing-chronic (one ear endpoint) and chronic-chronic, and sorted by |phi| within group.***

#### Panel A. Primary all-ages network (N = 18,939; 62 edges).

| Node 1 | Node 2 | phi | P value | Edge type |
|---|---|---|---|---|
| **Objective hearing loss** | Subjective hearing difficulty | 0.232 | 1.1e-223 | Hearing-internal |
| **Objective hearing loss** | Tinnitus | 0.217 | 5.9e-196 | Hearing-internal |
| **Subjective hearing difficulty** | Tinnitus | 0.179 | 5.5e-134 | Hearing-internal |
| **Subjective hearing difficulty** | Arthritis | 0.221 | 3.6e-203 | Hearing-chronic |
| **Subjective hearing difficulty** | Hypertension | 0.190 | 1.0e-150 | Hearing-chronic |
| **Tinnitus** | Arthritis | 0.182 | 1.9e-138 | Hearing-chronic |
| **Tinnitus** | Depression | 0.176 | 1.3e-129 | Hearing-chronic |
| **Tinnitus** | Hypertension | 0.160 | 1.9e-107 | Hearing-chronic |
| **Tinnitus** | Chronic bronchitis | 0.142 | 4.8e-85 | Hearing-chronic |
| **Subjective hearing difficulty** | Coronary heart disease | 0.132 | 9.6e-74 | Hearing-chronic |
| **Objective hearing loss** | Hypertension | 0.128 | 1.9e-69 | Hearing-chronic |
| **Subjective hearing difficulty** | Myocardial infarction | 0.126 | 2.3e-67 | Hearing-chronic |
| **Objective hearing loss** | Arthritis | 0.125 | 2.5e-66 | Hearing-chronic |
| **Subjective hearing difficulty** | Heart failure | 0.125 | 2.5e-66 | Hearing-chronic |
| **Subjective hearing difficulty** | Diabetes | 0.120 | 2.9e-61 | Hearing-chronic |
| **Subjective hearing difficulty** | Stroke | 0.112 | 1.3e-53 | Hearing-chronic |
| **Objective hearing loss** | Diabetes | 0.110 | 9.1e-52 | Hearing-chronic |
| **Subjective hearing difficulty** | Depression | 0.108 | 5.7e-50 | Hearing-chronic |
| **Subjective hearing difficulty** | Emphysema | 0.108 | 5.7e-50 | Hearing-chronic |
| **Coronary heart disease** | Myocardial infarction | 0.478 | <1e-300 | Chronic-chronic |
| **Hypertension** | Arthritis | 0.410 | <1e-300 | Chronic-chronic |
| **Coronary heart disease** | Heart failure | 0.377 | <1e-300 | Chronic-chronic |
| **Myocardial infarction** | Heart failure | 0.375 | <1e-300 | Chronic-chronic |
| **Diabetes** | Hypertension | 0.339 | <1e-300 | Chronic-chronic |
| **Diabetes** | Arthritis | 0.257 | 5.3e-274 | Chronic-chronic |
| **Chronic bronchitis** | Emphysema | 0.225 | 1.6e-210 | Chronic-chronic |
| **Arthritis** | Thyroid disorder | 0.224 | 1.2e-208 | Chronic-chronic |
| **Hypertension** | Heart failure | 0.220 | 2.4e-201 | Chronic-chronic |
| **Hypertension** | Myocardial infarction | 0.218 | 9.5e-198 | Chronic-chronic |
| **Hypertension** | Coronary heart disease | 0.213 | 7.1e-189 | Chronic-chronic |
| **Chronic bronchitis** | Arthritis | 0.203 | 9.6e-172 | Chronic-chronic |
| **Stroke** | Hypertension | 0.194 | 5.0e-157 | Chronic-chronic |
| **Hypertension** | Thyroid disorder | 0.188 | 1.4e-147 | Chronic-chronic |
| **Asthma** | Chronic bronchitis | 0.188 | 1.4e-147 | Chronic-chronic |
| **Heart failure** | Arthritis | 0.180 | 1.8e-135 | Chronic-chronic |
| **Diabetes** | Heart failure | 0.180 | 1.8e-135 | Chronic-chronic |
| **Coronary heart disease** | Arthritis | 0.179 | 5.5e-134 | Chronic-chronic |
| **Myocardial infarction** | Arthritis | 0.178 | 1.6e-132 | Chronic-chronic |
| **Stroke** | Myocardial infarction | 0.171 | 1.9e-122 | Chronic-chronic |
| **Stroke** | Heart failure | 0.168 | 2.9e-118 | Chronic-chronic |
| **Stroke** | Coronary heart disease | 0.167 | 7.0e-117 | Chronic-chronic |

| | | | | |
|---|---|---|---|---|
| **Diabetes** | Coronary heart disease | 0.164 | 8.6e-113 | Chronic-chronic |
| **Diabetes** | Myocardial infarction | 0.161 | 9.0e-109 | Chronic-chronic |
| **Depression** | Arthritis | 0.160 | 1.9e-107 | Chronic-chronic |
| **Stroke** | Arthritis | 0.159 | 3.9e-106 | Chronic-chronic |
| **Hypertension** | Chronic bronchitis | 0.149 | 1.9e-93 | Chronic-chronic |
| **Emphysema** | Arthritis | 0.148 | 3.2e-92 | Chronic-chronic |
| **Hypertension** | Liver disorder | 0.137 | 2.7e-79 | Chronic-chronic |
| **Depression** | Chronic bronchitis | 0.132 | 9.6e-74 | Chronic-chronic |
| **Arthritis** | Liver disorder | 0.131 | 1.2e-72 | Chronic-chronic |
| **Heart failure** | Emphysema | 0.129 | 1.6e-70 | Chronic-chronic |
| **Diabetes** | Thyroid disorder | 0.129 | 1.6e-70 | Chronic-chronic |
| **Stroke** | Diabetes | 0.129 | 1.6e-70 | Chronic-chronic |
| **Coronary heart disease** | Emphysema | 0.129 | 1.6e-70 | Chronic-chronic |
| **Diabetes** | Liver disorder | 0.123 | 2.8e-64 | Chronic-chronic |
| **Depression** | Hypertension | 0.121 | 2.9e-62 | Chronic-chronic |
| **Diabetes** | Chronic bronchitis | 0.120 | 2.9e-61 | Chronic-chronic |
| **Hypertension** | Emphysema | 0.117 | 2.5e-58 | Chronic-chronic |
| **Chronic bronchitis** | Liver disorder | 0.111 | 1.1e-52 | Chronic-chronic |
| **Myocardial infarction** | Emphysema | 0.110 | 9.1e-52 | Chronic-chronic |
| **Depression** | Liver disorder | 0.109 | 7.3e-51 | Chronic-chronic |
| **Heart failure** | Chronic bronchitis | 0.101 | 6.4e-44 | Chronic-chronic |

**Panel B. Adult (≥20 y) sensitivity network (N = 11,279; 44 edges).**

| **Node 1** | **Node 2** | **phi** | **P value** | **Edge type** |
|---|---|---|---|---|
| **Objective hearing loss** | Subjective hearing difficulty | 0.217 | 1.6e-117 | Hearing-internal |
| **Objective hearing loss** | Tinnitus | 0.187 | 9.1e-88 | Hearing-internal |
| **Subjective hearing difficulty** | Tinnitus | 0.150 | 3.9e-57 | Hearing-internal |
| **Subjective hearing difficulty** | Arthritis | 0.180 | 1.8e-81 | Hearing-chronic |
| **Tinnitus** | Depression | 0.150 | 3.9e-57 | Hearing-chronic |
| **Subjective hearing difficulty** | Hypertension | 0.137 | 5.9e-48 | Hearing-chronic |
| **Subjective hearing difficulty** | Coronary heart disease | 0.116 | 7.1e-35 | Hearing-chronic |
| **Tinnitus** | Arthritis | 0.112 | 1.3e-32 | Hearing-chronic |
| **Tinnitus** | Chronic bronchitis | 0.111 | 4.5e-32 | Hearing-chronic |
| **Subjective hearing difficulty** | Heart failure | 0.110 | 1.6e-31 | Hearing-chronic |
| **Subjective hearing difficulty** | Myocardial infarction | 0.109 | 5.4e-31 | Hearing-chronic |
| **Coronary heart disease** | Myocardial infarction | 0.469 | <1e-300 | Chronic-chronic |
| **Coronary heart disease** | Heart failure | 0.367 | <1e-300 | Chronic-chronic |
| **Myocardial infarction** | Heart failure | 0.365 | <1e-300 | Chronic-chronic |
| **Hypertension** | Arthritis | 0.313 | 2.7e-242 | Chronic-chronic |
| **Diabetes** | Hypertension | 0.274 | 3.6e-186 | Chronic-chronic |
| **Asthma** | Chronic bronchitis | 0.255 | 1.6e-161 | Chronic-chronic |
| **Chronic bronchitis** | Emphysema | 0.215 | 2.1e-115 | Chronic-chronic |
| **Diabetes** | Arthritis | 0.192 | 2.0e-92 | Chronic-chronic |
| **Hypertension** | Heart failure | 0.190 | 1.5e-90 | Chronic-chronic |
| **Hypertension** | Myocardial infarction | 0.183 | 3.9e-84 | Chronic-chronic |
| **Hypertension** | Coronary heart disease | 0.178 | 1.1e-79 | Chronic-chronic |
| **Arthritis** | Thyroid disorder | 0.166 | 1.5e-69 | Chronic-chronic |
| **Chronic bronchitis** | Arthritis | 0.163 | 3.9e-67 | Chronic-chronic |
| **Stroke** | Hypertension | 0.157 | 2.0e-62 | Chronic-chronic |
| **Stroke** | Myocardial infarction | 0.157 | 2.0e-62 | Chronic-chronic |
| **Diabetes** | Heart failure | 0.157 | 2.0e-62 | Chronic-chronic |
| **Stroke** | Heart failure | 0.155 | 6.9e-61 | Chronic-chronic |
| **Stroke** | Coronary heart disease | 0.154 | 4.0e-60 | Chronic-chronic |
| **Heart failure** | Arthritis | 0.148 | 1.1e-55 | Chronic-chronic |
| **Coronary heart disease** | Arthritis | 0.145 | 1.7e-53 | Chronic-chronic |
| **Myocardial infarction** | Arthritis | 0.143 | 4.3e-52 | Chronic-chronic |
| **Diabetes** | Coronary heart disease | 0.139 | 2.6e-49 | Chronic-chronic |
| **Diabetes** | Myocardial infarction | 0.136 | 2.8e-47 | Chronic-chronic |
| **Emphysema** | Arthritis | 0.125 | 3.2e-40 | Chronic-chronic |
| **Stroke** | Arthritis | 0.124 | 1.3e-39 | Chronic-chronic |
| **Asthma** | Emphysema | 0.120 | 3.4e-37 | Chronic-chronic |
| **Heart failure** | Emphysema | 0.120 | 3.4e-37 | Chronic-chronic |

| | | | | |
|---|---|---|---|---|
| **Coronary heart disease** | Emphysema | 0.119 | 1.3e-36 | Chronic-chronic |
| **Depression** | Arthritis | 0.118 | 5.0e-36 | Chronic-chronic |
| **Hypertension** | Thyroid disorder | 0.114 | 9.7e-34 | Chronic-chronic |
| **Depression** | Chronic bronchitis | 0.114 | 9.7e-34 | Chronic-chronic |
| **Stroke** | Diabetes | 0.103 | 7.5e-28 | Chronic-chronic |
| **Asthma** | Arthritis | 0.101 | 7.6e-27 | Chronic-chronic |

***Nesting of the two panels: 42 of the 44 adult edges are also present in the all-ages network; the 2 exceptions (asthma–emphysema, arthritis–asthma) fall just below the phi threshold in the all-ages cohort. Nesting against the complete-case network (eTable 4): intersection 48, union 64, Jaccard = 0.750, recall = 0.960. Machine-readable copies are in the Open Science Framework repository (https://osf.io/9cy4f/) under Source_Data/ (eTable5_edgelist_panelA_allages.csv, eTable5_edgelist_panelB_adult20.csv, eTable5_edgelists.json).***

**eTable 6. Adult (≥20 y) Sensitivity — SEQN-correct alignment**

| Node | Degree (full cohort, N=18,939) | Degree (adult ≥20, N=11,279) |
|---|---|---|
| **HL_OBJ** | 5 | 2 |
| **HL_SUB** | 11 | 7 |
| **TIN** | 6 | 5 |
| **ASTHMA** | 1 | 3 |
| **DEP** | 6 | 3 |
| **STR** | 7 | 6 |
| **DM** | 11 | 6 |
| **HTN** | 14 | 8 |
| **CHD** | 8 | 8 |
| **MI** | 8 | 7 |
| **CHF** | 9 | 8 |
| **BRON** | 9 | 5 |
| **EMP** | 7 | 5 |
| **ART** | 14 | 13 |
| **THY** | 3 | 2 |
| **LIV** | 5 | 0 |

*Note: recomputed from SEQN-correct merge; supersedes the prior positional version. Edge count: full 62 vs adult 44.*

**eTable 7. Sixteen-node ear-indexed comorbidity network — structural descriptors (NHANES, N = 18,939; phi > 0.10 & P < .05)**

| Node | System / community group | Weighted prevalence, non-missing, % | Degree | Chronic-disease degree | Betweenness (Freeman) | Community (greedy modularity) |
|---|---|---|---|---|---|---|
| **Objective hearing loss** | Ear | 8.0 | 5 | 3 | 0.002 | ear-depression |
| **Subjective hearing difficulty** | Ear | 6.1 | 11 | 9 | 0.050 | ear-depression |
| **Tinnitus** | Ear | 10.6 | 6 | 4 | 0.010 | ear-depression |
| **Hypertension** | Cardio-metabolic | 30.2 | 14 | 11 | 0.136 | metabolic-musculoskeletal |
| **Diabetes** | Cardio-metabolic | 7.6 | 11 | 9 | 0.075 | metabolic-musculoskeletal |
| **Coronary heart disease** | Cardio-metabolic | 3.2 | 8 | 7 | 0.003 | cardio-cerebrovascular |
| **Myocardial** | Cardio- | 3.3 | 8 | 7 | 0.003 | cardio- |

| | | | | | | |
|---|---|---|---|---|---|---|
| **infarction** | metabolic | | | | | cerebrovascular |
| **Heart failure** | Cardio-metabolic | 2.6 | 9 | 8 | 0.018 | cardio-cerebrovascular |
| **Stroke** | Cardio-cerebrovascular | 2.8 | 7 | 6 | 0.000 | cardio-cerebrovascular |
| **Asthma** | Respiratory | 15.7 | 1 | 1 | 0.000 | respiratory |
| **Chronic bronchitis** | Respiratory | 5.7 | 9 | 8 | 0.156 | respiratory |
| **Emphysema** | Respiratory | 1.9 | 7 | 6 | 0.010 | respiratory |
| **Arthritis** | Other chronic | 24.8 | 14 | 11 | 0.136 | metabolic-musculoskeletal |
| **Thyroid disorder** | Other chronic | 11.3 | 3 | 3 | 0.000 | metabolic-musculoskeletal |
| **Liver disorder** | Other chronic | 3.7 | 5 | 5 | 0.002 | metabolic-musculoskeletal |
| **Depression** | Other chronic | 10.8 | 6 | 4 | 0.007 | ear-depression |

*Note: prevalence is the weighted prevalence among respondents with non-missing data for that item, matching the first prevalence column of eTable 1. Betweenness is Freeman-normalised on the unweighted edge set, as in eTable 1 and Figure 1. 'Chronic-disease degree' = number of edges to non-ear nodes. Communities are from greedy modularity maximisation on the phi-weighted graph (4 communities, Q = 0.162); Louvain at the same resolution returns an identical partition.*

**eTable 8. Bidirectional two-sample Mendelian randomization — ear phenotypes <-> brain outcomes (24 IVW pairs; Bonferroni alpha = 0.0021)**

| Exposure | Outcome | Direction | nSNP | beta (95% CI) | P | Interpretation |
|---|---|---|---|---|---|---|
| **Objective HL** | Alzheimer's (FinnGen) | forward | 3 | -0.069 (-0.455, +0.318) | 0.73 | Null after Bonferroni |
| **Objective HL** | Cognition | forward | 3 | -0.008 (-0.070, +0.053) | 0.79 | Null after Bonferroni |
| **Objective HL** | Dementia | forward | 3 | +0.213 (-0.080, +0.507) | 0.15 | Null after Bonferroni |
| **Objective HL** | Stroke | forward | 3 | -0.087 (-0.466, +0.293) | 0.65 | Null after Bonferroni |
| **Subjective HL** | Alzheimer's (ieu-a-297) | forward | 22 | -0.571 (-1.595, +0.453) | 0.27 | Null after Bonferroni |
| **Subjective HL** | Alzheimer's (FinnGen) | forward | 22 | -0.229 (-1.727, +1.269) | 0.76 | Null after Bonferroni |
| **Subjective HL** | Cognition | forward | 22 | +0.053 (-0.225, +0.331) | 0.71 | Null after Bonferroni |
| **Subjective HL** | Dementia | forward | 22 | -0.195 (-1.383, +0.993) | 0.75 | Null after Bonferroni |
| **Subjective HL** | Stroke | forward | 23 | -0.407 (-1.118, +0.304) | 0.26 | Null after Bonferroni |
| **Alzheimer's** | Objective | reverse | 15 | -0.012 (- | 0.34 | Null after |

| | | | | | | |
|---|---|---|---|---|---|---|
| **(ieu-a-297)** | HL | | | 0.037, +0.013) | | Bonferroni |
| **Alzheimer's (ieu-a-297)** | Subjective HL | reverse | 14 | +0.001 (-0.001, +0.004) | 0.34 | Null after Bonferroni |
| **Alzheimer's (ieu-a-297)** | Tinnitus | reverse | 14 | +0.058 (-0.006, +0.122) | 0.077 | Null after Bonferroni |
| **Alzheimer's (FinnGen)** | Objective HL | reverse | 3 | +0.003 (-0.019, +0.025) | 0.81 | Null after Bonferroni |
| **Alzheimer's (FinnGen)** | Subjective HL | reverse | 3 | +0.001 (-0.001, +0.003) | 0.52 | Null after Bonferroni |
| **Alzheimer's (FinnGen)** | Tinnitus | reverse | 3 | +0.028 (-0.102, +0.158) | 0.68 | Null after Bonferroni |
| **Cognition** | Objective HL | reverse | 146 | -0.146 (-0.262, -0.031) | 0.013 | Null after Bonferroni |
| **Cognition** | Subjective HL | reverse | 146 | -0.007 (-0.020, +0.006) | 0.28 | Null after Bonferroni |
| **Cognition** | Tinnitus | reverse | 146 | +0.242 (-0.039, +0.523) | 0.091 | Null after Bonferroni |
| **Dementia** | Objective HL | reverse | 6 | +0.004 (-0.024, +0.032) | 0.79 | Null after Bonferroni |
| **Dementia** | Subjective HL | reverse | 6 | +0.001 (-0.002, +0.005) | 0.42 | Null after Bonferroni |
| **Dementia** | Tinnitus | reverse | 6 | +0.035 (-0.088, +0.158) | 0.58 | Null after Bonferroni |
| **Stroke** | Objective HL | reverse | 17 | -0.035 (-0.156, +0.087) | 0.58 | Null after Bonferroni |
| **Stroke** | Subjective HL | reverse | 17 | -0.001 (-0.023, +0.022) | 0.96 | Null after Bonferroni |
| **Stroke** | Tinnitus | reverse | 17 | +0.042 (-0.268, +0.351) | 0.79 | Null after Bonferroni |

*Note: IVW estimates; all 24 pairs null after Bonferroni correction. Full MR (MR-Egger, weighted median) with heterogeneity/pleiotropy diagnostics are in eTable 2-3.*

**eTable 9. CHARLS Longitudinal Models — Hearing Difficulty and Cognitive Decline (Panel A, Cox proportional hazards; Panel B, linear mixed model; Panel C, Kaplan-Meier survival)**

*Panel A. Cox proportional hazards (full model: hearing difficulty at 2011 → incident memory problem at 2013–2018; baseline prevalent cases excluded).*

| Exposure | Hazard | 95% CI | 95% CI | P | N | Events |
|---|---|---|---|---|---|---|

| | ratio | low | high | | | |
|---|---|---|---|---|---|---|
| **Hearing difficulty (vs normal)** | 1.571 | 1.27 | 1.94 | 4.0e-05 | 17,101 | 658 |

*Panel B. Linear mixed model (random slope; 0–30 composite cognition score). Specification: cog ~ hear + t + hear×t + age + C(sex) + C(edu); groups=id; REML.*

| Parameter | Coefficient | 95% CI low | 95% CI high | P / note |
|---|---|---|---|---|
| **Hearing (hear)** | -0.8039 | -1.0705 | -0.5374 | main effect |
| **Hearing × Time** | -0.0626 | -0.1439 | 0.0187 | P=0.131 |
| **7-y cognition change — normal** | -0.39 | | | estimated |
| **7-y cognition change — hearing** | -0.83 | | | estimated |
| **Sensitivity: random intercept (0–20 recall)** | -0.067 | | | P=0.001 |
| **7-y change — normal (sens.)** | -0.11 | | | |
| **7-y change — hearing (sens.)** | -0.58 | | | |

*Panel C. Kaplan–Meier 7-year survival probability by hearing status (for Fig. 4 reference).*

| Time (y) | Normal surv. | Hearing surv. | Normal n | Hearing n | Events (hearing) |
|---|---|---|---|---|---|
| **0** | 1.0000 | 1.0000 | 15636 | 1465 | 112 |
| **2** | 0.9948 | 0.9911 | 15636 | 1465 | 112 |
| **4** | 0.9856 | 0.9720 | 15636 | 1465 | 112 |
| **7** | 0.9651 | 0.9235 | 15636 | 1465 | 112 |

*Source: CHARLS 2011–2018 (n_baseline=17,596; hearing prevalence 8.92%). Cox and LMM numeric results are reproducible from the Open Science Framework repository (https://osf.io/9cy4f/), script charls_longitudinal_v2.py.*

### eFigure Captions

eFigure 1. Community-coloured ear-indexed comorbidity network (NHANES 2011-2012 + 2015-2016, N = 18,939). Nodes are coloured by greedy-modularity community on the phi-weighted graph (4 communities, Q = 0.162); edge width is proportional to the phi coefficient. The three ear nodes fall in a single ear-depression community together with depression; within that community objective hearing loss carries only 3 chronic-disease edges against 9 for subjective hearing difficulty, which is the empirical anchor of the audiometric-silence asymmetry.

eFigure 2. Single-cycle (2011-2012, G) cognitive subnet. Cognition (CFDCSR<=3) shown in relation to ear indicators and cardiometabolic nodes; illustrates the G-cycle-limited cognitive measurement and motivates the MR/CHARLS causal extension.

### eMethods. Supplementary Methods Detail

**MR instrument selection.** Genome-wide significant SNPs (P<5e-8) for each exposure were extracted from IEU OpenGWAS, clumped at r2<0.001 within 10 Mb (ieugwasr/TwoSampleMR), and retained only if F>10. Exposures: objective hearing loss (ebi-a-GCST90018857, 3 SNPs), subjective difficulty (ukb-a-257, 22-23 SNPs), tinnitus (ukb-b-14254, 1 SNP). Outcomes: cognition (ebi-a-GCST006572), Alzheimer disease (ieu-a-297; finn-b-G6_ALZHEIMER), dementia (finn-b-F5_DEMENTIA), stroke (ebi-a-GCST005838).

**LDSC genetic correlation.** LDSC (Bulik-Sullivan et al., Nat Genet 2015) estimated pairwise genetic correlation (rg) between ear exposures and brain outcomes using publicly available summary statistics; 6 estimable pairs, all |rg|<=0.12, all P>.19.

**Two-step MR.** Leg A tested ear exposure -> inflammatory mediators (CRP, ebi-a-GCST90029070; IL-6, ebi-a-GCST90012005); Leg B tested mediator -> Alzheimer disease as a positive control (CRP->AD IVW beta=-0.72, P=3e-8).

**CAUSE.** CAUSE latent-confounder model (v1.2.0) was specified for ear->brain pairs; estimation did not converge and is reported as non-estimable, excluded from primary triangulation.

**CHARLS analysis.** Baseline (2011) self-reported hearing difficulty (8.9% of 17,101) predicting incident self-reported memory problems (2011->2018) via Cox proportional hazards adjusted for age, sex, education. Cognitive trajectory via linear mixed-effects models. Scale-derived impairment cutoffs excluded as outcome.

**Reporting.** STROBE and STROBE-MR checklists completed (see submission package). Pre-registration: OSF 9cy4f (secondary-data preregistration, Part A retrospective; Transparent Changes A-D for post-registration tiers).
